# Distributed genetic architecture across the hippocampal formation implies common neuropathology across major brain disorders

**DOI:** 10.1101/2021.08.18.21262223

**Authors:** Shahram Bahrami, Kaja Nordengen, Alexey A. Shadrin, Oleksandr Frei, Dennis van der Meer, Anders M. Dale, Lars T. Westlye, Ole A. Andreassen, Tobias Kaufmann

**Affiliations:** Norwegian Centre for Mental Disorders Research (NORMENT), Division of Mental Health and Addiction, Oslo University Hospital & Institute of Clinical Medicine, University of Oslo, Oslo, Norway; School of Mental Health and Neuroscience, Faculty of Health, Medicine and Life Sciences, Maastricht University, Maastricht, The Netherlands; Department of Radiology, School of Medicine, University of California, San Diego, CA, USA; Department of Neurosciences, University of California San Diego, La Jolla, CA 92037, USA; Center for Multimodal Imaging and Genetics, University of California at San Diego, La Jolla, CA, 92037, USA; Department of Psychology, University of Oslo, Oslo, Norway; Department of Psychiatry and Psychotherapy, Tübingen Center for Mental Health (TüCMH), University of Tübingen, Tübingen, Germany

## Abstract

Despite its major role in complex human behaviours across the lifespan, including memory formation and decline, navigation and emotions, much of the genetic architecture of the hippocampal formation is currently unexplored. Here, through multivariate genome-wide association analysis in volumetric data from 35,411 individuals, we revealed 173 unique genetic loci with distributed associations across the hippocampal formation. We identified profound genetic overlap with eight major developmental and degenerative brain disorders, where common genes suggest partly age- and disorder-independent mechanisms underlying hippocampal pathology.

---

The hippocampal formation on each side of the medial temporal lobes of the brain plays critical roles in spatial and episodic memory^1,2^, navigation^3,4^, emotions^5^, and other complex human behaviours. Consequently, impaired or lesion-induced loss of hippocampal functioning has tremendous and diverse impact on cognitive and emotional functions^6,7^, and the hippocampus has been extensively studied across a wide variety of diseases and traits, including trajectories of development and aging.

While previous volumetric magnetic resonance imaging (MRI) studies of the human hippocampus have primarily investigated volume of the structure as a whole, recent advancements into deriving its subdivisions through adaptive segmentation has allowed for a more fine-grained assessment across multiple subregions^8^. The hippocampal formation comprises the histologically distinguishable subfields of the hippocampus proper as well as the dentate gyrus with its own subfields, and the neocortical subiculum, presubiculum and parasubiculum. In all but the latter, the hippocampal formation is also divided into an anterior (head) and a posterior part (body). The various contributions to the function of the hippocampal formation differ along the septo-temporal axis, where for example the anterior part is known to contribute to anxiety-related behaviours and cost-benefit decision making while the posterior part is particularly involved in spatial processing^5^. Such a dichotomy between the hippocampal head and the hippocampal body is also evident from patterns of neurogenesis. For example, although neurogenesis continues throughout life in the dentate gyrus^9^, it is especially the process in its anterior part that has been linked to emotion regulation and appears of relevance for the behavioural effects of antidepressants^10^. A total of 19 subregion volumes can currently be segmented with MRI^8^.

The rate of hippocampal maturation and change throughout the lifespan may capture information relevant to the study of major psychiatric and neurological disorders, where the age at which individual neurophysiological trajectories diverge from the norm may reflect key characteristics of the underlying pathophysiology^11^. Indeed, the hippocampal formation is known to be involved in disorders with typical onset during neurodevelopment^12^, such as autism spectrum disorders (ASD), attention-deficit hyperactivity disorder (ADHD), schizophrenia (SCZ) and bipolar disorder (BIP), through its roles in perception, memory processes, modulation of executive function, emotion regulation, among others^13-15^. Furthermore, through its involvement in stress response, the hippocampal formation is potentially involved in migraine (MIG)^16^ and tests of recollection memory indicate hippocampal dysfunction in major depression (MD)^17^, both of which are disorders that can appear at any stage from adolescence to old age. Finally, the hippocampal formation is implied in diseases that primarily emerge during senescence such as Parkinson’s disease (PD) and Alzheimer’s disease (AD). Emerging data suggests a complex hippocampal crosstalk among the dopaminergic and other transmitter systems in PD, involved in adaptive memory and motivated behaviour^18^. Loss of hippocampal functions like navigation and episodic memory are core markers of AD and hippocampal atrophy is an established finding^19,20^. Taken together, the hippocampal formation is a central and inter-connected structure that plays important roles in complex human behaviours and is implicated in a range of major psychiatric and neurological disorders across the lifespan.

The recent decade has brought significant progress towards a characterization of the genetic architecture of the hippocampal formation, from experimental manual mapping in mice^21^ to brain imaging based genome-wide-association studies (GWAS) in humans^22-24^, revealing fifteen independent loci when studying individual hippocampal subfields^24^. Given the broad functional portfolio of the hippocampal formation, however, it is clear that much of the genetic architecture remains to be explored, calling for further studies and novel analytical approaches^25^. Recent work revealed a distributed genetic architecture of human brain anatomy^26,27^ and function^28^ and suggested that capitalizing on this distributed nature in a multivariate GWAS approach can significantly improve the discovery beyond standard GWAS approaches^26^. We hypothesized that the genetic architecture within the hippocampal formation is distributed across its subregions and thus aimed to gain novel insights into the genetics of the hippocampal formation by deploying such multivariate GWAS approach. Further, given hippocampal involvement in major brain disorders across the lifespan, we used common neurological and psychiatric disorders as proxies for potential age-dependent hippocampal pathology, aiming to reveal gene variants potentially involving the hippocampus at different stages in life.

We accessed raw T1-weighted MRI data from 35,411 genotyped white British individuals (age range: aged 45 – 82 years, mean: 64.4 years, sd: 7.5 years, 51.7% females) from the UK Biobank^29^ (permission no. 27412) and segmented the hippocampal formation into 19 subregions in addition to total hippocampus volume (sum of all subfields) using FreeSurfer 7.1^8^. For each of these, we calculated the average volume between the left and right hemisphere and residualized for age, age squared, sex, scanning site, a proxy of image quality, intracranial volume and the first 20 genetic principal components. The resulting residuals were used in genetic analyses, feeding the 19 subregions alongside whole hippocampus volume into multivariate GWAS (MOSTest^26^).

## Multivariate approach identifies 173 loci associated with hippocampal formation

In line with our hypothesis, we found strong support of a distributed genetic architecture in the hippocampal formation. Multivariate GWAS revealed 173 unique genetic loci with distributed associations across the hippocampal formation. The upper panel of **Figure 1** depicts the corresponding multivariate statistics, highlighting the polygenic architecture of the hippocampal formation. For each of the 173 loci, the lower panel of **Figure 1** depicts statistics from univariate GWASs of individual hippocampal subregions. The elevated univariate statistics for multiple hippocampal subregions in some of the same loci supports a distributed genetic architecture across the hippocampal formation, which is also supported by genetic correlation analysis of the univariate GWASs of the individual subregions (**Supplementary Figure 1, Supplementary Table 1**). Whereas the strongest hits among our 173 discovered loci are also implied in univariate analysis, a large share of the 173 loci showed elevated yet not genome-wide significant effects at univariate level. By capitalizing on these distributed effects across subregions, the multivariate approach boosted discovery. **Supplementary Table 2** provides additional details on the 173 discovered loci, most of which were not identified in previous hippocampus GWAS. **Supplementary Figure 2** depicts corresponding quantile-quantile (Q–Q) plots, including one from permutation testing that confirms validity of the multivariate test statistic. **Supplementary Figure 3** covers additional validations, including a cross-ethnic replication attempt in an independent set of 5262 individuals with non-white ethnicity.

**Figure 1.**
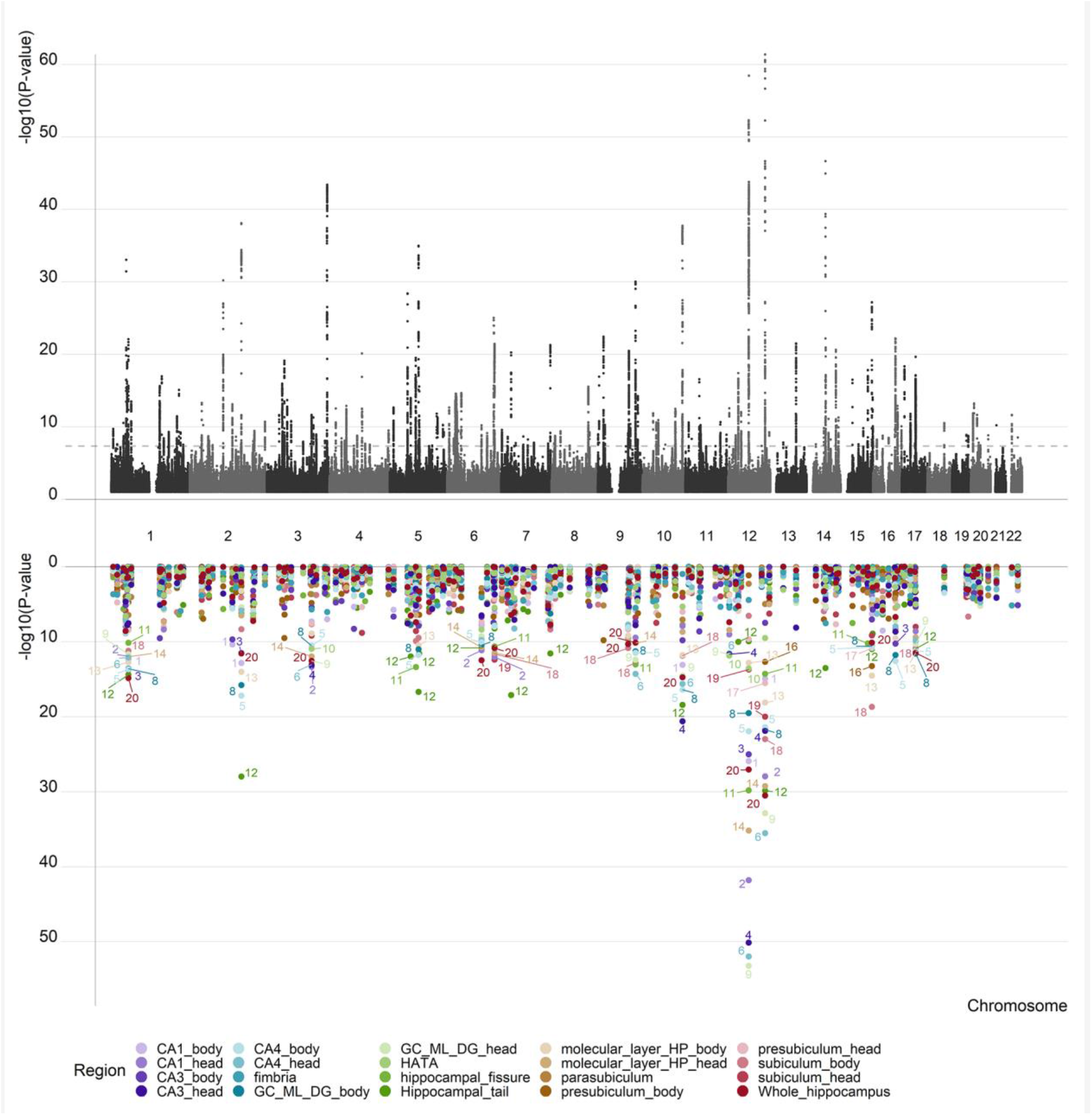
The multivariate framework discovered 173 independent loci significantly associated with the hippocampal formation. The upper panel illustrates the multivariate GWAS statistics for the entire formation with 173 significant loci. The lower panel for each of the 173 unique loci depicts the corresponding statistics from univariate GWASs of single subregions (one colour per subregion).

## Functional mapping and annotation identifies 69 genes robustly associated with hippocampal formation

We functionally annotated all candidate SNPs (n=21430) that were in linkage disequilibrium (r2 ≥ 0.6) with one of the independent significant SNPs using Functional Mapping and Annotation of GWAS (FUMA)^30^. About 90% of the SNPs had a minimum chromatin state of 1–7, thus suggesting they were in open chromatin regions^31,32^, and 5.6% were in regulatomeDB category 1 or 2, suggesting potential regulatory function^31^ (**Supplementary Figure 4A-B**). A majority of these SNPs were intronic (50.7%) or intergenic (29.9%) and 1.0% were exonic (**Supplementary Figure 4C**).

We mapped the 173 loci implied for the hippocampal formation to 813 genes based on different mapping strategies (positional, expression quantitative trait loci (eQTL), chromatin state mapping and MAGMA analysis). Out of these, 69 genes were identified by all four mapping strategies (**Figure 2**), supporting robustness of these findings. **Supplementary Tables 3-6** provide additional details. Among the genes mapped from loci with strongest GWAS effects we found *LEMD3* (also known as *MAN1*) and *RNFT2* (also known as *TMEM118*), both important for immune regulation^33,34^. Another strong hit was *FAM53B*, known for its important roles in neurodevelopment such as control of cell proliferation and maintenance of a pluripotent state^35^. Rs2151909 on chromosome 6 was mapped to six different genes by all four mapping strategies: *KATNA1, LATS1, NUP43, PCMT1, GINM1* and *LRP11 (*also known as *SORL1)*. The latter is a neuronal apolipoprotein E receptor important in pathogenesis of AD^36^.

**Figure 2.**
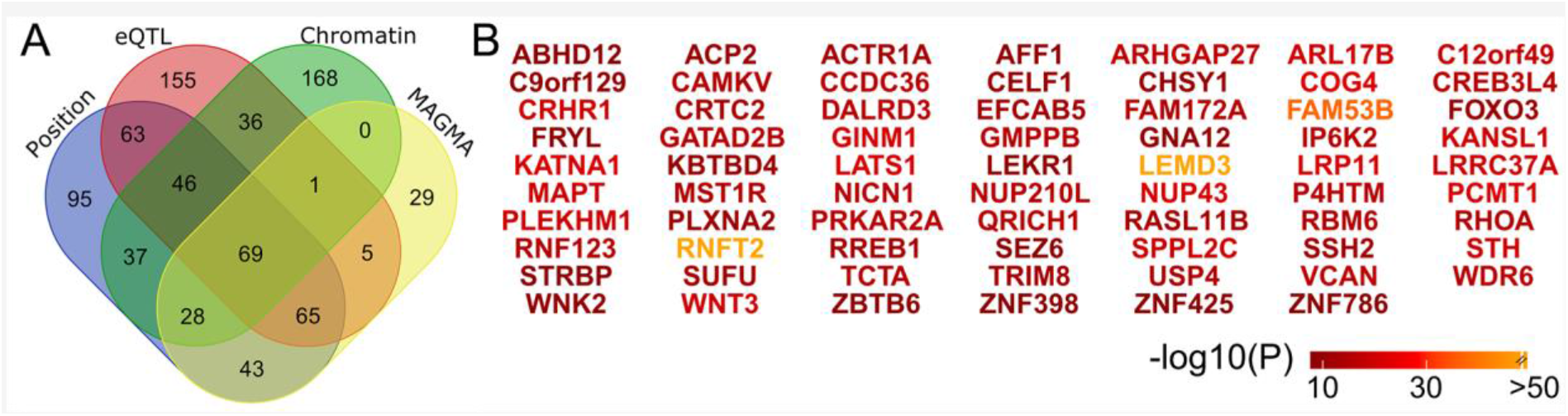
Gene mapping of the 173 loci associated with the hippocampal formation implied 69 genes by all mapping strategies. (A) Venn diagram showing number of genes mapped by the four different strategies. (B) The 69 genes implied by all four strategies with color-coded P-values.

Genome-wide gene-based association studies (GWGAS; two-sided P < 2.752 × 10^−6^) through MAGMA identified 240 unique genes across the hippocampus (**Supplementary Table 5**). Many of the significant gene sets reflected processes related to early development (**Supplementary Table 7**), such as neurogenesis (P_Bonf_ =6.8 × 10^−8^), regulation of anatomical structure morphogenesis (P_Bonf_ = 1.0 × 10^−7^) and neuronal differentiation (P_Bonf_ =5.0 × 10^−7^), potentially indicating that individual differences in hippocampal volumes later in life may be largely determined early in development. Also, when focusing on the 69 genes implicated from all four mapping strategies (see above), the gene-sets reflect processes related to early development like regulation of cell morphogenesis (P = 4.4 × 10^−6^ and P = 1.2 × 10^−5^) and axogenesis (P = 2.3 × 10^−5^) but also to biological processes on reproduction, like multi organism reproductive process (P = 2.2 × 10^−5^), sexual reproduction (P = 3.0 × 10^−5^) and gamete generation (P = 5.0 × 10^−5^), which might potentially represent a later hormonal modulation of hippocampal neurogenesis (**Supplementary Table 8**). Although pathways important for neural differentiation are overrepresented (**Supplementary Table 9**) we also found dominant pathways representing processes prominent during the lifetime, like damage mechanisms (spinal cord injury and DNA damage response) and inflammation (*IL-5* signaling pathway, *IL-2* signaling pathway and putative anti-inflammatory metabolites formation from EPA) in addition to overrepresented pathways on vitamin B, D and E metabolism.

## Genetic overlap between hippocampal formation and common brain disorders

Using major neurological and psychiatric disorders as proxies for potential age-dependent hippocampal pathology, we studied the genetic overlap between hippocampal formation and eight disorders - ASD, ADHD, SCZ, BIP, MIG, MD, PD and AD – with typical onset times across the lifespan. The commonly used approach - genetic correlations of the disorders with individual hippocampus subregions - did not show significant associations after Bonferroni correction for multiple comparisons (**Supplementary Table 10**). However, conditional Q–Q plots^37^ conditioning the multivariate statistic of hippocampal formation on the disorders and vice versa showed a clear pattern of pleiotropic enrichment in both directions (**Supplementary Figure 5**). Conjunctional FDR analysis^37,38^ allowed us to test for shared loci between the hippocampus and each of the disorders. Strikingly, we identified 7 loci significantly (conjFDR<0.05) overlapping with ADHD, 4 loci with ASD, 63 with BIP, 130 with SCZ, 23 with MD, 81 with MIG, 24 with AD and 13 loci significantly overlapping with PD (**Figure 3A**).

**Figure 3.**
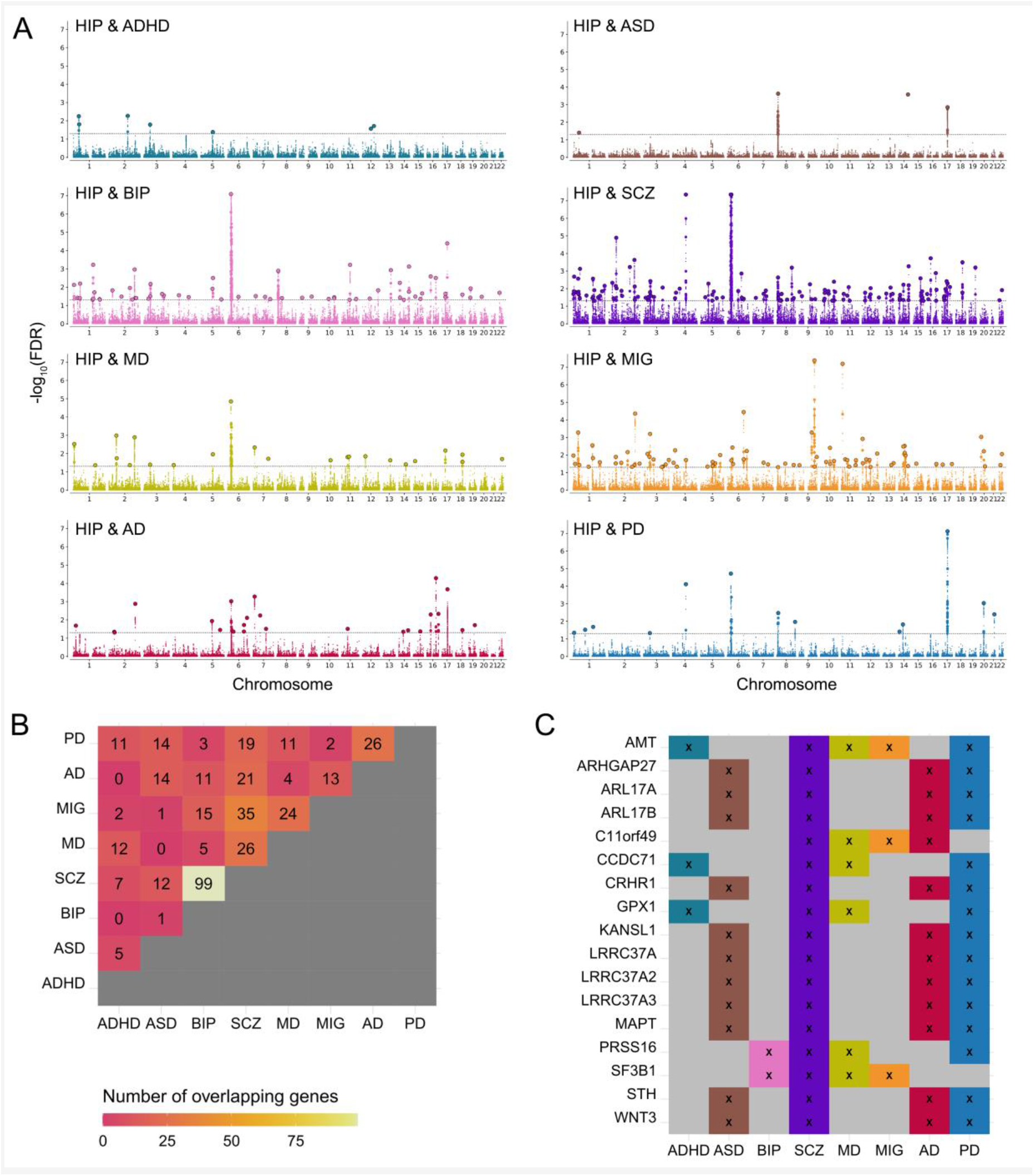
Genetic overlap between hippocampal formation and eight disorders with different onset times as proxies of age-dependent hippocampal pathology. (A) For each disorder, a conjunctional FDR Manhattan plot is shown, illustrating the –log10 transformed conjunctional FDR values for each SNP on the y axis and chromosomal positions along the x axis. The dotted horizontal line represents the threshold for significant shared associations (conjFDR < 0.05). Independent lead SNPs are encircled in black. (B) Various genes mapped from the conjunctional FDR analysis were implied to overlap between hippocampal formation and multiple disorders. The figure illustrates the total number of genes implied in each combination of disorders. For example, 99 of the genes overlapping between hippocampal formation and SCZ were also found to overlap between hippocampal formation and BIP. (C) Panel C complements panel B with a list of genes that were implied for more than 3 disorders. One gene was mapped for five disorders (*AMT*) and 16 genes were mapped for four disorders. ASD autism spectrum disorder, ADHD attention deficit hyperactivity disorder, SCZ schizophrenia, BIP bipolar disorder, MIG migraine, MD, major depression, PD Parkinson’s disease, AD Alzheimer’s disease and HIP hippocampal formation.

**Supplementary Tables 11-18** provide a full list of loci overlapping between hippocampal formation and the disorders. We mapped each of these loci to genes using positional, eQTL and chromatin state mapping (**Supplementary Table 19)** and checked for genes that were implicated for multiple disorders. By far strongest overlap was found between SCZ and BIP, where 99 of the genes overlapping between hippocampal formation and SCZ were also found to overlap between hippocampal formation and BIP (**Figure 3B**). While this overlap may be expected given the relatedness of the disorders, it is particularly noteworthy that we found large overlap between other combinations of disorders as well, some of which pertain very different onset times across the lifespan such as ASD and AD (14 genes), ADHD and PD (11 genes), or ASD and PD (14 genes). Many genes were implied for more than two disorders, and **Figure 3C** depicts the subset implicated as overlapping with hippocampal formation for at least four disorders. Again, it is particular worth noting the co-occurance for distinct disorders in different phases of life. For example, the most frequently mapped gene was the *AMT* gene involved in glycinergic neurotransmission, found to overlap between hippocampal formation and ADHD, SCZ, MD, MIG and PD, respectively (**Supplementary Table 19**). Other examples are the tau protein associated genes *MAPT* and *STH*, found for ASD, SCZ, AD and PD, or the *GPX1* gene, known to protect cells from oxidative stress and here found for ADHD, SCZ, MD and PD. This may illustrate genetic mechanisms independent of life phases and may suggest that some of the pleiotropy between brain disorders might be explained by shared mechanisms in hippocampal pathology.

## Discussion

Taken together, our multivariate GWAS of the volumes of the hippocampal formation revealed a plethora of genomic loci not identified in previous work, implicating a distributed nature of effects on hippocampus. The mapped genes have roles in neurobiological processes across the lifespan. Importantly, the profound overlap between hippocampal formation and common brain disorders and the identification of some of the same genes implied for disorders with onset in different phases of life suggests age-independent neuropathology and pinpoints potential disease-independent drug targets.

The distributed genetic architecture across the hippocampal formation, here revealed through 173 hippocampus-associated loci and 69 genes mapped by four mapping strategies, pointed at pathways with involvement across the lifespan, starting with embryogenic brain development, like axon guidance and neuronal migration, then involving neuronal plasticity processes including angiogenesis, and finally neurodegenerative processes such as DNA repair. We also found overrepresented pathways on vitamin B, D and E metabolism, supporting their role in neuropathology^39^. Finally, the identified immune-related pathways support that inflammatory responses could be involved in hippocampal pathology mediating brain disease^40-42^.

Our results also suggest that several genes have a role in hippocampal pathology across multiple brain disorders, with onset times ranging across the lifespan. These findings not only support the notion of pleiotropy across a spectrum of neurological and psychiatric disorders but may also pinpoint to both, age- and disorder-independent drug targets. For example, our discovery of the *AMT* gene overlapping between hippocampal formation and ADHD, SCZ, MD, MIG and PD, respectively, may implicate an age-independent role of glycine in hippocampal pathology. The *AMT* gene codes for aminomethyltransferase (T-protein), an enzyme crucial for the glycine decarboxylase complex (GCS) in mitochondria. Glycine is a primary inhibitory neurotransmitter in the spinal cord and brainstem, but increasing evidence show important glycine involvement also in the hippocampal formation^43,44^. Glycine exerts a tonic inhibitory role through extrasynaptic glycine receptor chloride channels^45^, in addition to modulation of NMDA receptors in the hippocampal formation^46,47^. Through regulation of both glycine and serine synthesis and cleavage, aminomethyltransferase as part of the glycine decarboxylase complex, may provide a homeostatic regulation of hippocampal function and plasticity by simultaneous activation of excitatory NMDA receptors and inhibitory glycine receptors^43,48^. The glycine site on the NMDA receptors is currently under investigation as a promising drug target for several of the disorders that we here associated with the *AMT* gene, including ADHD^49,50^, PD^51^, SCZ^52^ and MD^53^, either by direct glycine supplementation, other substances working on the same receptor site or by increasing endogenous glycine by inhibiting the glycine transporter. Another example for a potential age- and disorder-independent drug target is the microtubule-associated protein tau (*MAPT*) gene, which was here implicated for ASD, SCZ, AD and PD. Indeed, tau has for long been a marker of AD and PD^54^ yet has recently also gained focus for ASD^55^, with animal models suggesting that tau reduction may prevent behavioural signs of this neurodevelopmental disorder^56^. Taken together, our results therefore add support for disorder-independent gene targets for hippocampal pathology across the lifespan, including *AMT* and *MAPT*, among others, and illustrate how the multivariate GWAS approach can reveal overlapping biochemical mechanisms underlying different disorders and traits.

In conclusion, our results suggest a polygenic architecture of the hippocampal formation, distributed across its subregions. The genetic overlap with various developmental and degenerative brain disorders implicated genes that may be relevant targets for future studies into the mechanisms underlying hippocampal functioning and pathology across the lifespan. With several of the findings fitting currently studied treatment targets (e.g. the glycine site on the NMDA receptor), our results also confirm the utility of the approach and suggest that capitalizing on the distributed nature of genetic effects on the brain will be instrumental in our future endeavours to further understand mechanisms underlying the brain and its disorders.

## Data Availability

In this study we used brain imaging and genetics data from the UK Biobank [https://www.ukbiobank.ac.uk/], and GWAS summary statistics obtained from the Psychiatric Genomics Consortium [https://www.med.unc.edu/pgc/], 23andMe [https://www.23andme.com/], International headache genetics Consortium (IHGC) [http://www.headachegenetics.org], the International Genomics of Alzheimer's Project [http://web.pasteur-lille.fr/en/recherche/u744/igap/igap_download.php], and the International Parkinson Disease Genomics Consortium [https://pdgenetics.org/]. The latter included 23andMe data, which was made available through 23andMe under an agreement with 23andMe that protects the privacy of the 23andMe participants [https://research.23andme.com/collaborate/#dataset-access/].
The summary statistics for hippocampal formation derived in this study are made available in our github repository [https://github.com/norment/open-science] upon acceptance of the manuscript for publication. Likewise, we will publish the FUMA results on the FUMA website [https://fuma.ctglab.nl/browse/] upon acceptance.

## Acknowledgments

The authors were funded by the Research Council of Norway (TK: 276082. OAA: 213837, 223273, 248778, 273291, 262656, 229129, 283798, 311993. LTW: 204966, 249795, 273345), the South-Eastern Norway Regional Health Authority (OAA: 2013-123, 2017-112, 2019-108. LTW: 2014-097, 2015-073, 2016-083), Norwegian Health Association (SB: 22731), Stiftelsen Kristian Gerhard Jebsen, the European Research Council (LTW: ERCStG 802998), and an NVIDIA Corporation GPU Grant (TK). The funding bodies had no role in the analysis or interpretation of the data; the preparation, review or approval of the manuscript; nor in the decision to submit the manuscript for publication. This work was performed on the TSD (Tjeneste for Sensitive Data) facilities, owned by the University of Oslo, operated and developed by the TSD service group at the University of Oslo, IT-Department (USIT) and on resources provided by UNINETT Sigma2 - the National Infrastructure for High Performance Computing and Data Storage in Norway. The research has been conducted using the UK Biobank Resource (access code 27412) and using summary statistics for various brain disorders that partly included 23andMe data. We would like to thank the research participants and employees of UK Biobank, the 23andMe and the consortia contributing summary statistics for making this work possible.

## Author contributions

SB and TK conceived the study and analysed the data. KN and TK interpreted the results and spearheaded the writing. SB and TK drafted the online methods. All authors gave conceptual input on the methods and/or results and all authors contributed to and approved the final manuscript.

## Online Methods

### Sample and pre-processing of imaging and genetic data

The UK Biobank was approved by the National Health Service National Research Ethics Service (ref. 11/NW/0382). We accessed raw T1-weighted magnetic resonance brain imaging data from 35,411 genotyped white British from the UK Biobank^29^ (age range: aged 45 – 82 years, mean: 64.4 years, sd: 7.5 years, 51.7% females) for the main analysis, and of 5262 individuals with non-white ethnicity (age range: 45 - 81, mean: 62.9, sd: 7.6 years, 53.6% females) for the replication in independent data.

We processed T1-weighted images using the standard *recon-all* pipeline in Freesurfer 5.3^57^, and subsequently segmented the hippocampal formation using Freesurfer 7.1^8^. For genetic analyses, we followed the standard quality control procedures to the UK Biobank v3 imputed genetic data and removed SNPs with an imputation quality score < 0.5, a minor allele frequency < 0.05, missing in more than 5% of individuals, and failing the Hardy Weinberg equilibrium tests at a *P* < 1e-6.

### Multivariate genome-wide association analysis

For each of the 19 regions of the hippocampal formation – parasubiculum, presubiculum head and body, subiculum head and body, CA1/CA3/CA4 head and body, GC-ML-DG head and body, molecular layer head and body, HATA, fimbria, hippocampal tail, hippocampal fissure – as well as for total hippocampus volume we calculated the average volume between the left and right hemisphere and subsequently residualized the volumes for age, age squared, sex, scanning site, Euler number as a proxy of image quality, intracranial volume and the first 20 genetic principal components. The resulting residuals for the 20 regions were jointly fed into MOSTest^26^ analysis, yielding a multivariate GWAS summary statistic across all 20 features. For comparison to state-of-the art univariate approaches, we also performed univariate GWAS (extracted from the univariate stream of MOSTest^26^). Supplemental genetic correlation analyses were performed using LD-score regression^58,59^.

To identify genetic loci we uploaded this summary statistic to the FUMA platform v1.3.6^30^. Using the 1000GPhase3 EUR as reference panel, we identified independent significant SNPs at the statistical significance threshold *P* < 5e−8. All SNPs at r^2^<0.6 with each other were considered as *independent significant SNPs* and a fraction of the independent significant SNPs in approximate linkage equilibrium with each other at r^2^<0.1 were considered as *lead SNPs*. Post-GWAS eQTL analysis was also performed using FUMA. The full FUMA analysis including the corresponding parameter file will be made available through https://fuma.ctglab.nl/browse upon acceptance.

### Genetic overlap between hippocampal formation and major brain disorders

We accessed GWAS summary statistics for migraine (MIG) from International headache genetics Consortium^60^ and for Parkinson’s disease (PD) from the International Parkinson Disease Genomics Consortium^61,62^. The latter included 23andMe data. 23andMe participants provided informed consent and participated in the research online, under a protocol approved by the external AAHRPP-accredited IRB, Ethical & Independent Review Services (E&I Review). Furthermore, from the Psychiatric Genomics Consortium we accessed summary statistics for attention deficit hyperactivity disorder (ADHD)^63^, autism spectrum disorder (ASD)^64^, bipolar disorder (BIP)^65^ and major depression (MD)^66^. Finally, we included data from recent studies of schizophrenia (SCZ)^67^ and of Alzheimer’s disease (AD)^68^

Using conjunctional FDR statistics (FDR<0.05)^37,38^, we identified shared variants associated with hippocampal formation and each of the above-mentioned brain disorders. Two genomic regions, the extended major histocompatibility complex genes region (hg19 location Chr 6: 25119106– 33854733) and chromosome 8p23.1 (hg19 location Chr 8: 7242715–12483982) for all cases and *MAPT* region for PD and *APOE* region for AD and ASD, respectively were excluded from the FDR fitting procedures because complex correlations in regions with intricate LD can bias FDR estimation.

We submitted the results from conjunctional FDR to FUMA v1.3.6^30^ to annotate the genomic loci with conjFDR value<0.10 having an r2≥0.6 with one of the independent significant SNPs. FUMA annotates associated SNPs based on functional Categories, Combined Annotation Dependent Depletion (CADD) scores which predicts the deleteriousness of SNPs on protein structure/function^69^, RegulomeDB scores which predicts regulatory functions^31^; and chromatin states that shows the transcription/regulation effects of chromatin states at the SNP locus^70^. We also conducted Gene Ontology gene-set analyses based on FUMA’s gene ontology classification system^30^ and pathway analyses^71^ for the all mapped genes and the 69 common mapped genes of hippocampal formation.

## Data availability

In this study we used brain imaging and genetics data from the UK Biobank [https://www.ukbiobank.ac.uk/], and GWAS summary statistics obtained from the Psychiatric Genomics Consortium [https://www.med.unc.edu/pgc/], 23andMe [https://www.23andme.com/], International headache genetics Consortium (IHGC) [http://www.headachegenetics.org], the International Genomics of Alzheimer’s Project [http://web.pasteur-lille.fr/en/recherche/u744/igap/igap_download.php], and the International Parkinson Disease Genomics Consortium [https://pdgenetics.org/]. The latter included 23andMe data, which was made available through 23andMe under an agreement with 23andMe that protects the privacy of the 23andMe participants [https://research.23andme.com/collaborate/#dataset-access/].

The summary statistics for hippocampal formation derived in this study are made available in our github repository [https://github.com/norment/open-science] upon acceptance of the manuscript for publication. Likewise, we will publish the FUMA results on the FUMA website [https://fuma.ctglab.nl/browse/] upon acceptance.

## Code availability

All code and software needed to generate the results is available as part of public resources, specifically MOSTest (https://github.com/precimed/mostest), FUMA (https://fuma.ctglab.nl/), conjunctional FDR (https://github.com/precimed/pleiofdr/) and LD score regression (https://github.com/bulik/ldsc).

## Supplementary Figures

**Supplementary Figure 1.**
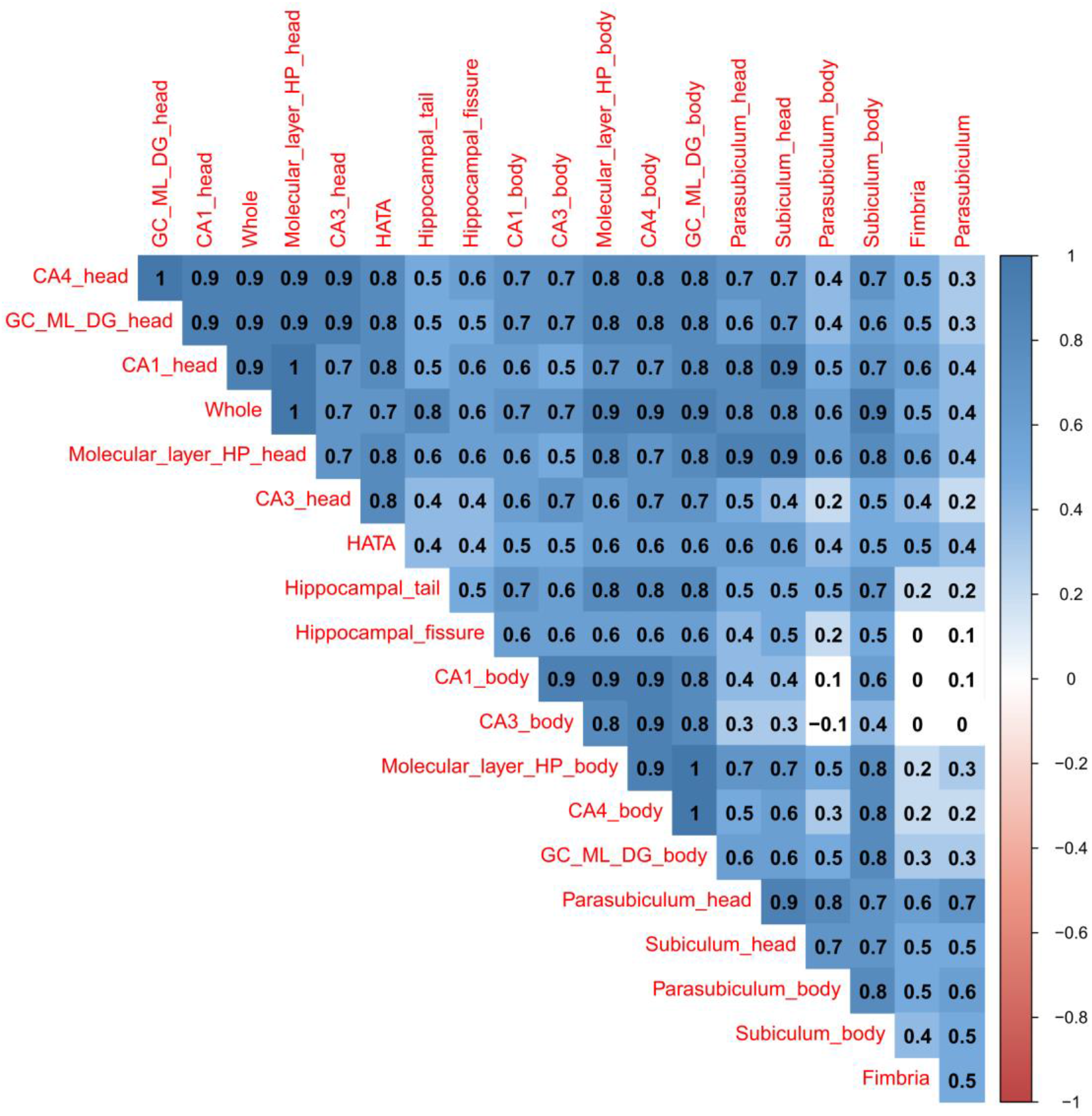
Genetic correlations between hippocampus volumes. For each pair of regions, the LD-score regression based genetic correlations of the univariate summary statistics are presented.

**Supplementary Figure 2.**
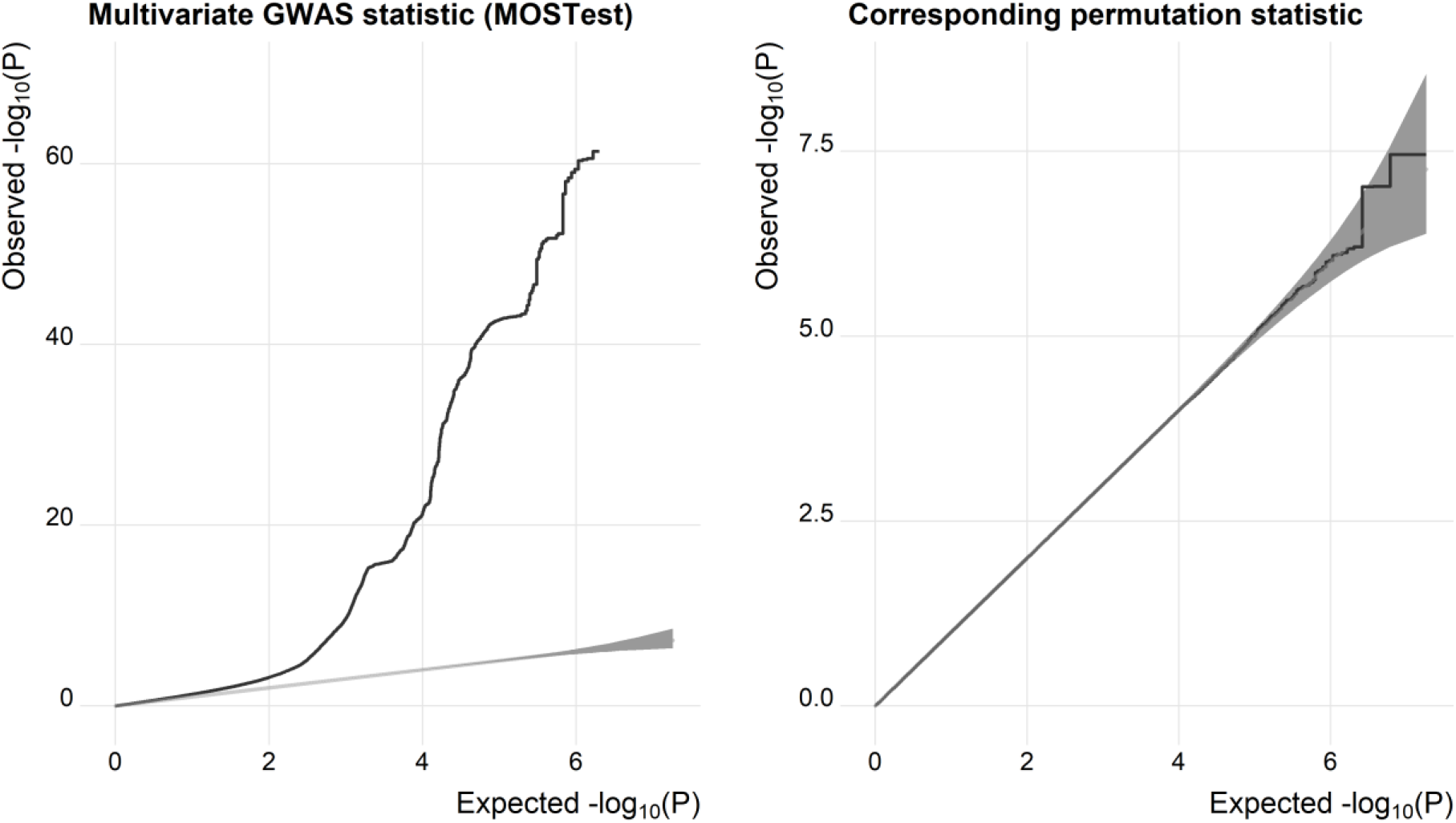
Quantile-quantile plots from MOSTest analysis. The left panel depicts signal from MOSTest analysis. The right panel shows test statistics under null (from permutation testing) and confirms validity of the MOSTest test statistics.

**Supplementary Figure 3.**
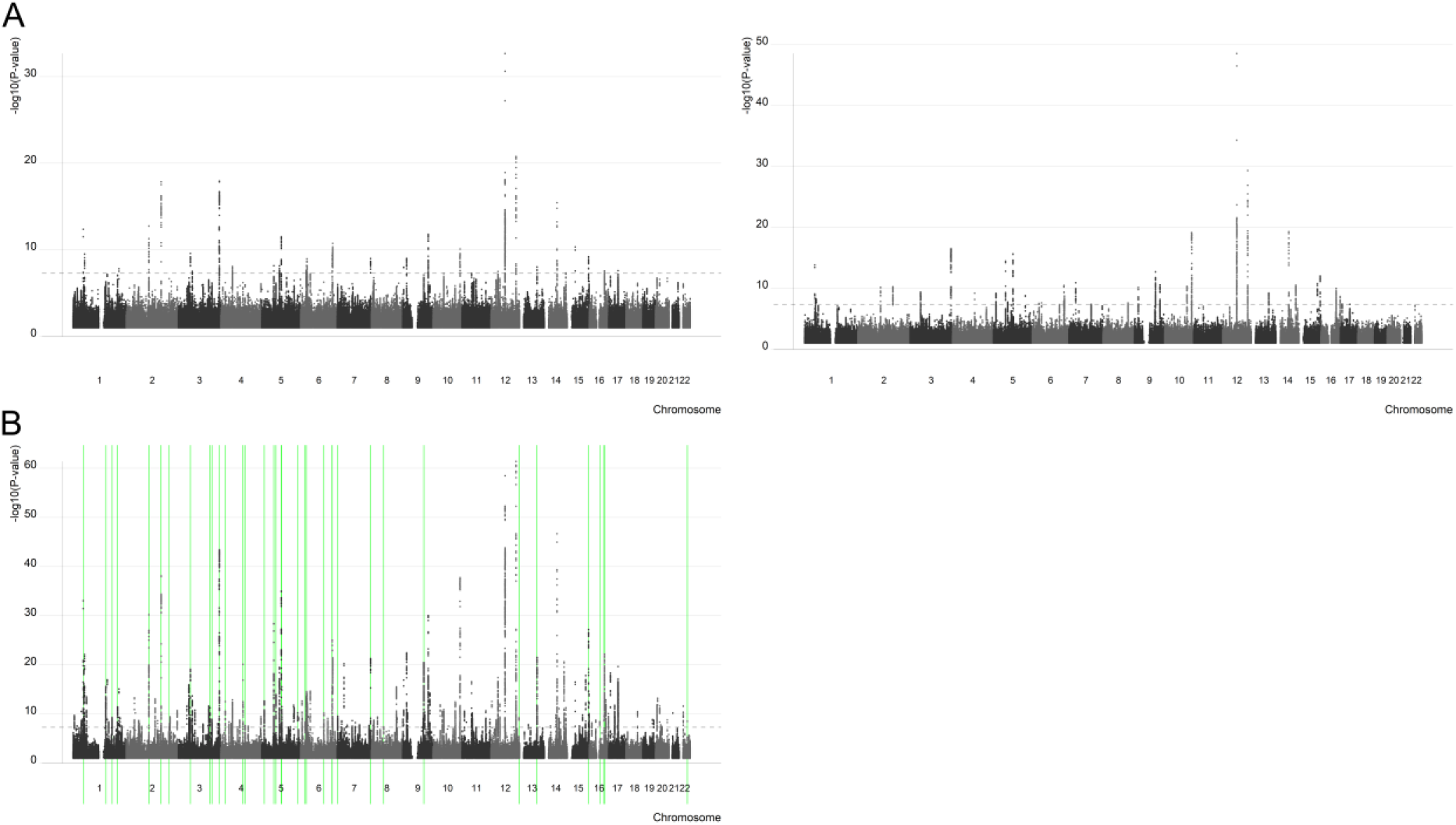
Validation and replication. We performed two analyses to further assess the robustness of our findings. A) We randomly split the sample used in the main analysis in half and used one half as the discovery sample and the other as the replication sample. We performed a MOSTest analysis in both samples (the figure shows the corresponding manhattan plots) and calculated replication rates as an indicator of robustness of the MOSTest approach. From this split half analysis we found that 96.9% of the loci replicated at nominal P and 46.9% replicated at 5e-8. B) We performed a cross-ethnic replication of the MOSTest analysis described in the main text using independent data from 5262 individuals with non-white ethnicity. 31.9% of the loci found in the full sample of white British individuals replicated at nominal P in the independent replication sample of non-white individuals (indicated with green lines). Given the small sample and differences in ethnicity, replication rates were fairly low yet met our level of expectation, in line with earlier work on other brain structures^27^.

**Supplementary Figure 4.**
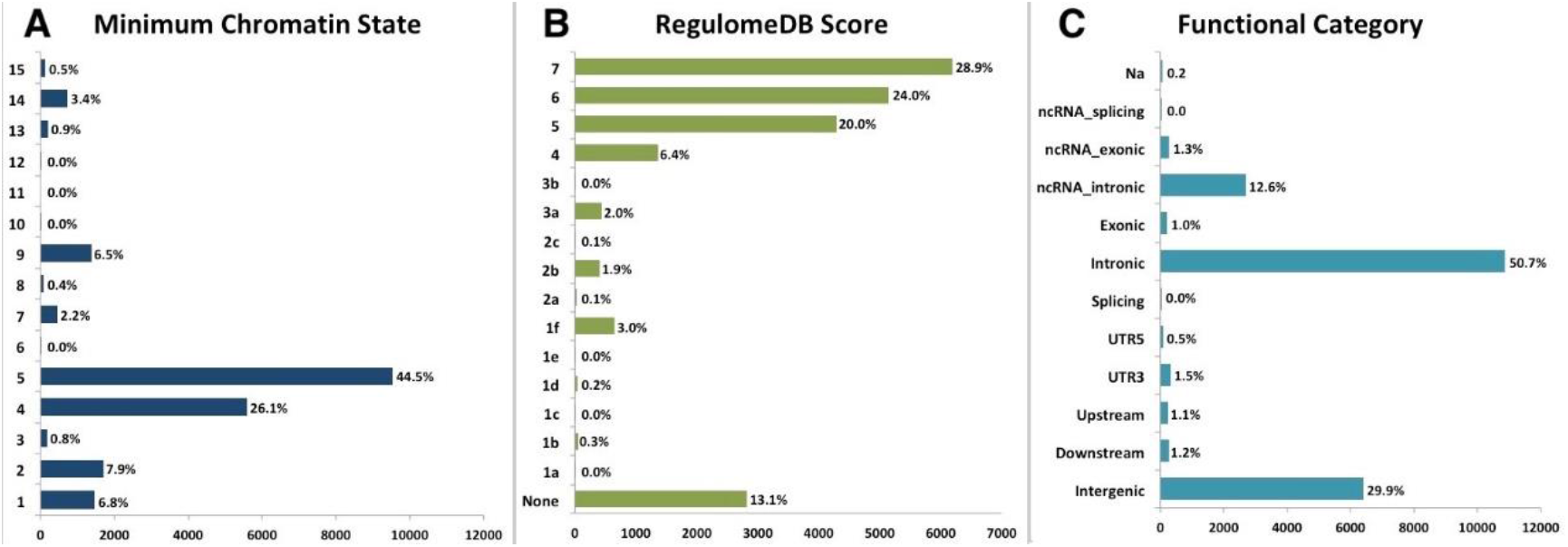
Distribution of the annotation for all SNPs in the significant genetic loci from the hippocampus GWAS including (A) the minimum chromatin state across 127 tissue and cell types for SNPs in the significant genomic loci, with lower states indicating higher accessibility and states 1–7 referring to open chromatin states, (B) the distribution of RegulomeDB scores for SNPs in the significant genomic loci, with a low score indicating a higher likelihood of having a regulatory function and (C) the distribution of functional consequences of SNPs in the significant genomic risk loci.

**Supplementary Figure 5.**
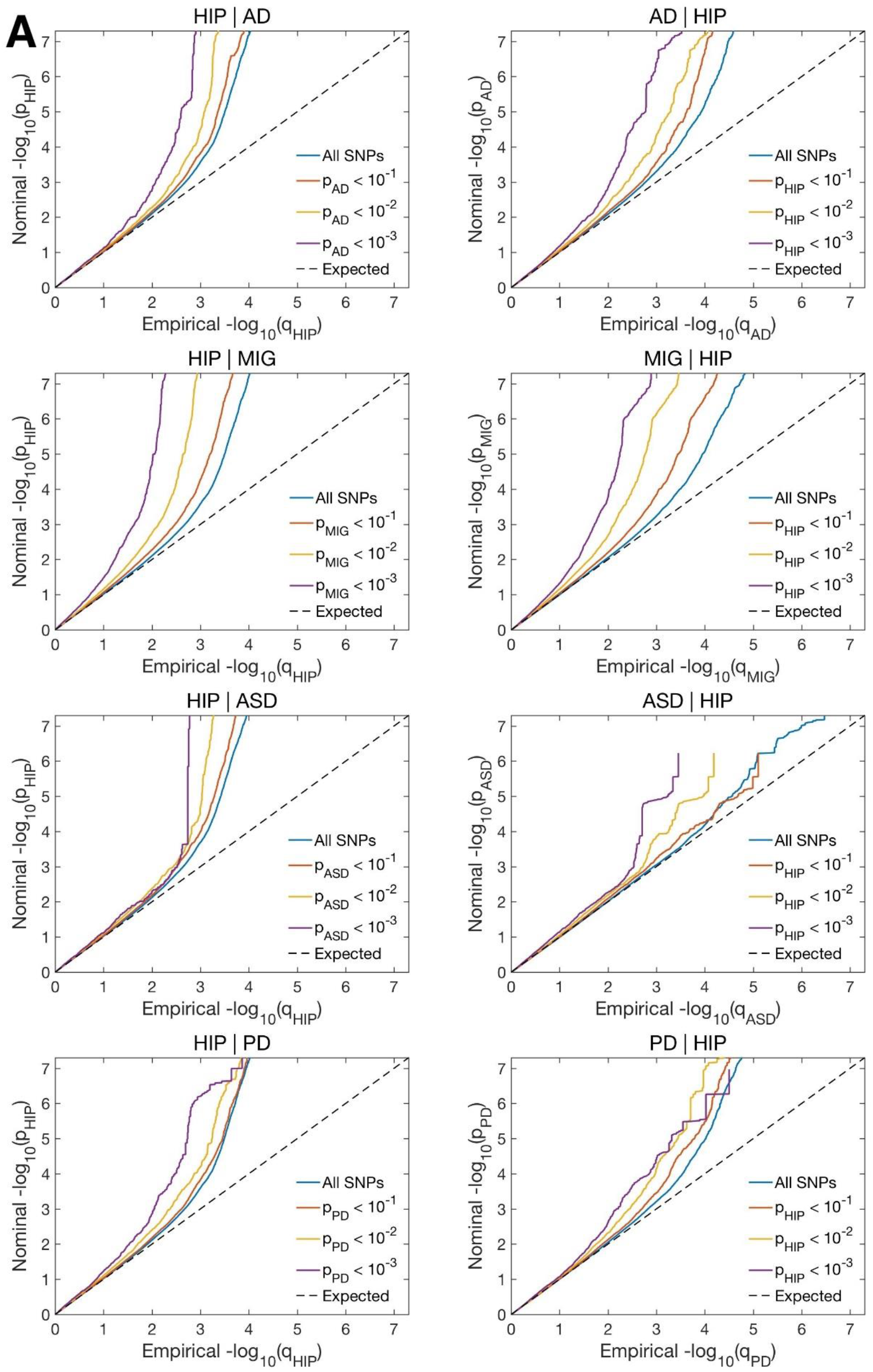
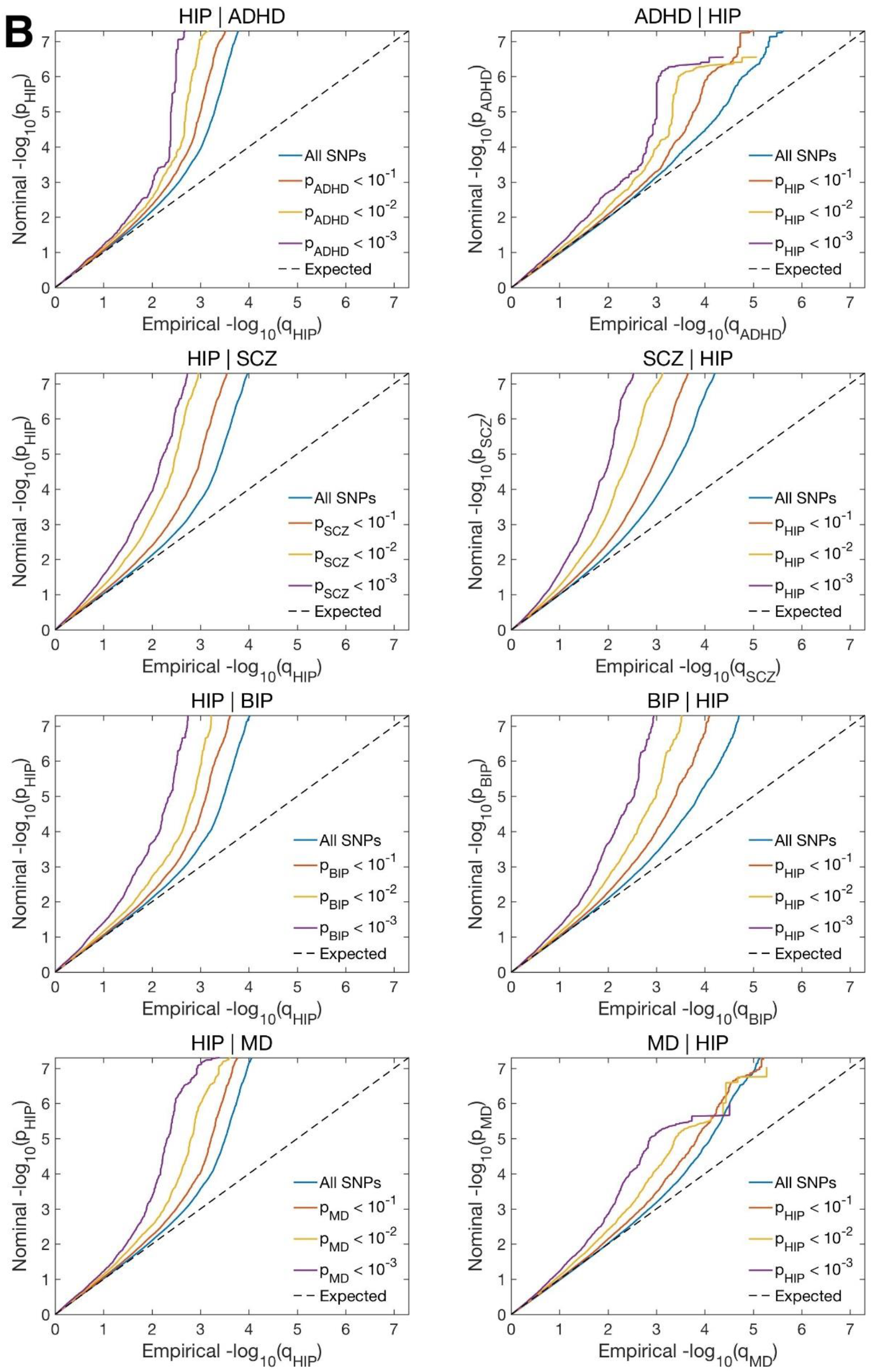
Conditional Q-Q plots for hippocampus given associations with the disorder (left figures) and vice versa (right figures). ADHD: attention deficit hyperactivity disorder. AD: Alzheimer’s disease. MIG: migraine. ASD: autism spectrum disorder. PD: Parkinson’s disease. SCZ: schizophrenia. BIP: bipolar disorder. MD: major depression.

## References

1 Burgess, N., Maguire, E. A. & O’Keefe, J. The human hippocampus and spatial and episodic memory. Neuron 35, 625–641, doi:10.1016/s0896-6273(02)00830-9 (2002).

2 Eichenbaum, H. A cortical-hippocampal system for declarative memory. Nat Rev Neurosci 1, 41–50, doi:10.1038/35036213 (2000).

3 Epstein, R. A., Patai, E. Z., Julian, J. B. & Spiers, H. J. The cognitive map in humans: spatial navigation and beyond. Nat Neurosci 20, 1504–1513, doi:10.1038/nn.4656 (2017).

4 Maguire, E. A. et al. Navigation-related structural change in the hippocampi of taxi drivers. Proc Natl Acad Sci U S A 97, 4398–4403, doi:10.1073/pnas.070039597 (2000).

5 Bannerman, D. M. et al. Hippocampal synaptic plasticity, spatial memory and anxiety. Nat Rev Neurosci 15, 181–192, doi:10.1038/nrn3677 (2014).

6 Chudasama, Y., Wright, K. S. & Murray, E. A. Hippocampal lesions in rhesus monkeys disrupt emotional responses but not reinforcer devaluation effects. Biol Psychiatry 63, 1084–1091, doi:10.1016/j.biopsych.2007.11.012 (2008).

7 Scoville, W. B. & Milner, B. Loss of recent memory after bilateral hippocampal lesions. J Neurol Neurosurg Psychiatry 20, 11–21, doi:10.1136/jnnp.20.1.11 (1957).

8 Iglesias, J. E. et al. A computational atlas of the hippocampal formation using ex vivo, ultra-high resolution MRI: Application to adaptive segmentation of in vivo MRI. Neuroimage 115, 117–137, doi:10.1016/j.neuroimage.2015.04.042 (2015).

9 Eriksson, P. S. et al. Neurogenesis in the adult human hippocampus. Nat Med 4, 1313–1317, doi:10.1038/3305 (1998).

10 Sahay, A. & Hen, R. Adult hippocampal neurogenesis in depression. Nat Neurosci 10, 1110–1115, doi:10.1038/nn1969 (2007).

11 Kaufmann, T. et al. Common brain disorders are associated with heritable patterns of apparent aging of the brain. Nat Neurosci 22, 1617–1623, doi:10.1038/s41593-019-0471-7 (2019).

12 Marin, O. Developmental timing and critical windows for the treatment of psychiatric disorders. Nat Med 22, 1229–1238, doi:10.1038/nm.4225 (2016).

13 Godsil, B. P., Kiss, J. P., Spedding, M. & Jay, T. M. The hippocampal-prefrontal pathway: the weak link in psychiatric disorders? Eur Neuropsychopharmacol 23, 1165–1181, doi:10.1016/j.euroneuro.2012.10.018 (2013).

14 Heckers, S. et al. Impaired recruitment of the hippocampus during conscious recollection in schizophrenia. Nat Neurosci 1, 318–323, doi:10.1038/1137 (1998).

15 Eastwood, S. L. & Harrison, P. J. Hippocampal synaptic pathology in schizophrenia, bipolar disorder and major depression: a study of complexin mRNAs. Mol Psychiatry 5, 425–432, doi:10.1038/sj.mp.4000741 (2000).

16 Borsook, D., Maleki, N., Becerra, L. & McEwen, B. Understanding migraine through the lens of maladaptive stress responses: a model disease of allostatic load. Neuron 73, 219–234, doi:10.1016/j.neuron.2012.01.001 (2012).

17 MacQueen, G. M. et al. Course of illness, hippocampal function, and hippocampal volume in major depression. Proc Natl Acad Sci U S A 100, 1387–1392, doi:10.1073/pnas.0337481100 (2003).

18 Calabresi, P., Castrioto, A., Di Filippo, M. & Picconi, B. New experimental and clinical links between the hippocampus and the dopaminergic system in Parkinson’s disease. Lancet Neurol 12, 811–821, doi:10.1016/S1474-4422(13)70118-2 (2013).

19 Masters, C. L. et al. Alzheimer’s disease. Nat Rev Dis Primers 1, 15056, doi:10.1038/nrdp.2015.56 (2015).

20 Barnes, J. et al. A meta-analysis of hippocampal atrophy rates in Alzheimer’s disease. Neurobiol Aging 30, 1711–1723, doi:10.1016/j.neurobiolaging.2008.01.010 (2009).

21 Thompson, C. L. et al. Genomic anatomy of the hippocampus. Neuron 60, 1010–1021, doi:10.1016/j.neuron.2008.12.008 (2008).

22 Hibar, D. P. et al. Novel genetic loci associated with hippocampal volume. Nat Commun 8, 13624, doi:10.1038/ncomms13624 (2017).

23 Stein, J. L. et al. Identification of common variants associated with human hippocampal and intracranial volumes. Nat Genet 44, 552–561, doi:10.1038/ng.2250 (2012).

24 van der Meer, D. et al. Brain scans from 21,297 individuals reveal the genetic architecture of hippocampal subfield volumes. Mol Psychiatry 25, 3053–3065, doi:10.1038/s41380-018-0262-7 (2020).

25 Vilor-Tejedor, N. et al. Genetic Influences on Hippocampal Subfields: An Emerging Area of Neuroscience Research. Neurol Genet 7, e591, doi:10.1212/NXG.0000000000000591 (2021).

26 van der Meer, D. et al. Understanding the genetic determinants of the brain with MOSTest. Nat Commun 11, 3512, doi:10.1038/s41467-020-17368-1 (2020).

27 van der Meer, D. et al. The genetic architecture of human cortical folding. bioRxiv, 2021.2001.2013.426555, doi:10.1101/2021.01.13.426555 (2021).

28 Roelfs, D. et al. Genetic overlap between multivariate measures of human functional brain connectivity and psychiatric disorders. medRxiv, 2021.2006.2015.21258954, doi:10.1101/2021.06.15.21258954 (2021).

29 Bycroft, C. et al. The UK Biobank resource with deep phenotyping and genomic data. Nature 562, 203–209, doi:10.1038/s41586-018-0579-z (2018).

30 Watanabe, K., Taskesen, E., van Bochoven, A. & Posthuma, D. Functional mapping and annotation of genetic associations with FUMA. Nat Commun 8, 1826, doi:10.1038/s41467-017-01261-5 (2017).

31 Boyle, A. P. et al. Annotation of functional variation in personal genomes using RegulomeDB. Genome Res 22, 1790–1797, doi:10.1101/gr.137323.112 (2012).

32 Roadmap Epigenomics, C. et al. Integrative analysis of 111 reference human epigenomes. Nature 518, 317–330, doi:10.1038/nature14248 (2015).

33 Hellemans, J. et al. Loss-of-function mutations in LEMD3 result in osteopoikilosis, Buschke-Ollendorff syndrome and melorheostosis. Nat Genet 36, 1213–1218, doi:10.1038/ng1453 (2004).

34 Tong, Y. et al. The RNFT2/IL-3Ralpha axis regulates IL-3 signaling and innate immunity. JCI Insight 5, doi:10.1172/jci.insight.133652 (2020).

35 Thermes, V. et al. Medaka simplet (FAM53B) belongs to a family of novel vertebrate genes controlling cell proliferation. Development 133, 1881–1890, doi:10.1242/dev.02350 (2006).

36 Cuccaro, M. L. et al. SORL1 mutations in early- and late-onset Alzheimer disease. Neurol Genet 2, e116, doi:10.1212/NXG.0000000000000116 (2016).

37 Andreassen, O. A. et al. Improved detection of common variants associated with schizophrenia and bipolar disorder using pleiotropy-informed conditional false discovery rate. PLoS Genet 9, e1003455, doi:10.1371/journal.pgen.1003455 (2013).

38 Smeland, O. B. et al. Discovery of shared genomic loci using the conditional false discovery rate approach. Hum Genet 139, 85–94, doi:10.1007/s00439-019-02060-2 (2020).

39 Eyles, D. W., Burne, T. H. & McGrath, J. J. Vitamin D, effects on brain development, adult brain function and the links between low levels of vitamin D and neuropsychiatric disease. Front Neuroendocrinol 34, 47–64, doi:10.1016/j.yfrne.2012.07.001 (2013).

40 Edvinsson, L., Haanes, K. A. & Warfvinge, K. Does inflammation have a role in migraine? Nat Rev Neurol 15, 483–490, doi:10.1038/s41582-019-0216-y (2019).

41 Nordengen, K. et al. Glial activation and inflammation along the Alzheimer’s disease continuum. J Neuroinflammation 16, 46, doi:10.1186/s12974-019-1399-2 (2019).

42 Yuan, N., Chen, Y., Xia, Y., Dai, J. & Liu, C. Inflammation-related biomarkers in major psychiatric disorders: a cross-disorder assessment of reproducibility and specificity in 43 meta-analyses. Transl Psychiatry 9, 233, doi:10.1038/s41398-019-0570-y (2019).

43 Keck, T., Lillis, K. P., Zhou, Y. D. & White, J. A. Frequency-dependent glycinergic inhibition modulates plasticity in hippocampus. J Neurosci 28, 7359–7369, doi:10.1523/JNEUROSCI.5618-07.2008 (2008).

44 Keck, T. & White, J. A. Glycinergic inhibition in the hippocampus. Rev Neurosci 20, 13–22, doi:10.1515/revneuro.2009.20.1.13 (2009).

45 Zhang, L. H., Gong, N., Fei, D., Xu, L. & Xu, T. L. Glycine uptake regulates hippocampal network activity via glycine receptor-mediated tonic inhibition. Neuropsychopharmacology 33, 701–711, doi:10.1038/sj.npp.1301449 (2008).

46 Johnson, J. W. & Ascher, P. Glycine potentiates the NMDA response in cultured mouse brain neurons. Nature 325, 529–531, doi:10.1038/325529a0 (1987).

47 Nong, Y. et al. Glycine binding primes NMDA receptor internalization. Nature 422, 302–307, doi:10.1038/nature01497 (2003).

48 Liu, L. et al. Role of NMDA receptor subtypes in governing the direction of hippocampal synaptic plasticity. Science 304, 1021–1024, doi:10.1126/science.1096615 (2004).

49 Hegvik, T. A. et al. Druggable genome in attention deficit/hyperactivity disorder and its co-morbid conditions. New avenues for treatment. Mol Psychiatry, doi:10.1038/s41380-019-0540-z (2019).

50 ClincalTrial. Sublingual Glycine vs. Placebo on Attentional Difficulties and Hyperactivity in Prepuberal Children, <https://ClinicalTrials.gov/show/NCT02655276> (Completed).

51 Heresco-Levy, U., Shoham, S. & Javitt, D. C. Glycine site agonists of the N-methyl-D-aspartate receptor and Parkinson’s disease: a hypothesis. Mov Disord 28, 419–424, doi:10.1002/mds.25306 (2013).

52 Heresco-Levy, U. et al. Efficacy of high-dose glycine in the treatment of enduring negative symptoms of schizophrenia. Arch Gen Psychiatry 56, 29–36, doi:10.1001/archpsyc.56.1.29 (1999).

53 Huang, C. C. et al. Inhibition of glycine transporter-I as a novel mechanism for the treatment of depression. Biol Psychiatry 74, 734–741, doi:10.1016/j.biopsych.2013.02.020 (2013).

54 Zhang, X. et al. Tau Pathology in Parkinson’s Disease. Front Neurol 9, 809, doi:10.3389/fneur.2018.00809 (2018).

55 Grigg, I. et al. Tauopathy in the young autistic brain: novel biomarker and therapeutic target. Transl Psychiatry 10, 228, doi:10.1038/s41398-020-00904-4 (2020).

56 Tai, C. et al. Tau Reduction Prevents Key Features of Autism in Mouse Models. Neuron 106, 421–437 e411, doi:10.1016/j.neuron.2020.01.038 (2020).

## References of the Online Methods

57 Fischl, B. et al. Whole brain segmentation: automated labeling of neuroanatomical structures in the human brain. Neuron 33, 341–355, doi:10.1016/s0896-6273(02)00569-x (2002).

58 Bulik-Sullivan, B. et al. An atlas of genetic correlations across human diseases and traits. Nat Genet 47, 1236–1241, doi:10.1038/ng.3406 (2015).

59 Bulik-Sullivan, B. K. et al. LD Score regression distinguishes confounding from polygenicity in genome-wide association studies. Nat Genet 47, 291–295, doi:10.1038/ng.3211 (2015).

60 Gormley, P. et al. Meta-analysis of 375,000 individuals identifies 38 susceptibility loci for migraine. Nat Genet 48, 856–866, doi:10.1038/ng.3598 (2016).

61 Nalls, M. A. et al. Large-scale meta-analysis of genome-wide association data identifies six new risk loci for Parkinson’s disease. Nat Genet 46, 989–993, doi:10.1038/ng.3043 (2014).

62 Chang, D. et al. A meta-analysis of genome-wide association studies identifies 17 new Parkinson’s disease risk loci. Nat Genet 49, 1511–1516, doi:10.1038/ng.3955 (2017).

63 Demontis, D. et al. Discovery of the first genome-wide significant risk loci for attention deficit/hyperactivity disorder. Nat Genet 51, 63–75, doi:10.1038/s41588-018-0269-7 (2019).

64 Grove, J. et al. Identification of common genetic risk variants for autism spectrum disorder. Nat Genet 51, 431–444, doi:10.1038/s41588-019-0344-8 (2019).

65 Mullins, N. et al. Genome-wide association study of more than 40,000 bipolar disorder cases provides new insights into the underlying biology. Nat Genet 53, 817–829, doi:10.1038/s41588-021-00857-4 (2021).

66 Wray, N. R. et al. Genome-wide association analyses identify 44 risk variants and refine the genetic architecture of major depression. Nat Genet 50, 668–681, doi:10.1038/s41588-018-0090-3 (2018).

67 Pardinas, A. F. et al. Common schizophrenia alleles are enriched in mutation-intolerant genes and in regions under strong background selection. Nat Genet 50, 381–389, doi:10.1038/s41588-018-0059-2 (2018).

68 Wightman, D. P. et al. Largest GWAS (N=1,126,563) of Alzheimer’s Disease Implicates Microglia and Immune Cells. medRxiv, 2020.2011.2020.20235275, doi:10.1101/2020.11.20.20235275 (2020).

69 Kircher, M. et al. A general framework for estimating the relative pathogenicity of human genetic variants. Nat Genet 46, 310–315, doi:10.1038/ng.2892 (2014).

70 Zhu, Z. et al. Integration of summary data from GWAS and eQTL studies predicts complex trait gene targets. Nat Genet 48, 481–487, doi:10.1038/ng.3538 (2016).

71 Kamburov, A., Wierling, C., Lehrach, H. & Herwig, R. ConsensusPathDB--a database for integrating human functional interaction networks. Nucleic Acids Res 37, D623–628, doi:10.1093/nar/gkn698 (2009).

